# Severe Periodontitis Biomarker Identification by Deep Salivary Proteome Profiling with Astral DIA Mass Spectrometry

**DOI:** 10.64898/2026.04.24.26351658

**Authors:** Xiaoyu Yu, Runting Yan, Hairui Li, Yu Xie, Mengning Bi, Yuan Li, Andrea Roccuzzo, Maurizio S. Tonetti

## Abstract

**Aim:** To comprehensively characterize the salivary proteome in periodontitis using Orbitrap Astral data-independent acquisition mass spectrometry (DIA-MS), identify an atlas of differentially expressed proteins (DEPs), and develop a machine learning-derived multi-protein biomarker panel for non-invasive diagnosis of stage III/IV periodontitis.

**Materials and Methods:** Unstimulated saliva samples from 199 participants (periodontal health/gingivitis, n=120; stage III/IV periodontitis, n=79) were analyzed by Orbitrap Astral DIA-MS. DEPs were identified, and pathway enrichment analysis was performed. A two-tier machine learning pipeline—integrating pathway-based feature selection with cross-validated evaluation—was applied to identify the optimal diagnostic panel.

**Results:** Orbitrap Astral DIA-MS quantified 5,597 salivary proteins and 1,966 DEPs (|log_2_FC|>0.5, FDR<0.05). Pathway analysis identified 14 periodontitis-relevant KEGG pathways, including Th17 cell differentiation, IL-17 signaling, neutrophil extracellular trap formation, and complement and coagulation cascades. A four-protein panel (TEC, RAC1, MAPK14, KRT17) achieved an area under the curve (AUC) of 0.985 ± 0.010, with 83% sensitivity and 100% specificity. The panel was corroborated using public datasets.

**Conclusions:** To our knowledge, this study represents the first application of Orbitrap Astral DIA mass spectrometry in periodontitis research, establishing a disease-specific DEPs atlas and a salivary biomarker panel with high diagnostic accuracy for stage III/IV periodontitis, providing a foundation for future external validation studies.

## 1. Introduction

Periodontitis is a chronic inflammatory disease characterized by progressive destruction of tooth-supporting tissues, with an estimated global prevalence of 11% in severe stages (Bernabe et al., 2020) and bidirectional associations with systemic conditions, including cardiovascular disease, diabetes mellitus, and adverse pregnancy outcomes (Blanco-Pintos et al., 2025; Sanz et al., 2020). Accurate diagnosis is essential for appropriate clinical management and prevention of further tissue destruction.

The gold standard for periodontitis diagnosis relies on clinical examination parameters, including probing pocket depth (PPD), clinical attachment level (CAL), and radiographic bone loss, as outlined in the 2017 Classification of Periodontal Diseases and Conditions and the recent Consensus on periodontal diagnosis (Herrera et al., 2025; Tonetti, Greenwell, & Kornman, 2018). These methods require professional examination with specialized instruments, limiting accessibility for population-level screening. The 2017 Classification acknowledged the potential utility of molecular biomarkers to enhance diagnostic accuracy, yet clinical translation remains limited (Herrera et al., 2025).

Saliva is an attractive diagnostic medium due to its non-invasive collection, ease of handling, and its rich composition of host-derived molecules that reflect local and systemic health status (Giannobile et al., 2009). Previous studies have identified candidate biomarkers, including matrix metalloproteinase-8 (MMP8), matrix metalloproteinase-9 (MMP9), interleukin-1β (IL-1β), and S100 proteins (S100A8, S100A9), with moderate to good diagnostic accuracy for single biomarkers, while multi-protein panels performed better (Arias-Bujanda et al., 2020; Blanco-Pintos et al., 2025; Grant et al., 2022). Among these, aMMP-8 point-of-care tests showed pooled sensitivity of only 0.59 (95% CrI: 0.42–0.75) with high false-negative rates across disease stages (Deng, Pelekos, Jin, & Tonetti, 2021; Li et al., 2025; Bi et al., 2025). Multi-protein panels have shown improved performance but remain constrained by limited proteome coverage (Bostanci et al., 2018; Grant et al., 2022) suggesting additional disease-relevant molecules remain to be discovered.

Mass spectrometry enables reproducible and comprehensive protein profiling (Ludwig et al., 2018). SWATH-MS has recently been applied to the salivary proteome in periodontitis (Blanco-Pintos et al., 2025), but proteome coverage on conventional DIA platforms remains limited. The Orbitrap Astral offers superior scan speed, sensitivity, and proteome depth, and has been applied to salivary proteomics in aging and obstructive sleep apnea, but not yet in periodontal disease research—representing a critical gap (Jiao et al., 2025; Lu et al., 2025).

Identifying differentially expressed proteins does not automatically establish diagnostic utility; rigorous predictive modeling and validation are essential for clinical translation (Gürsoy & Kantarci, 2022). As an initial step, this study focused on extreme-phenotype comparisons to maximize discriminatory power. Specifically, this study aimed to: (1) comprehensively profile the salivary proteome in periodontal health/gingivitis versus stage III/IV periodontitis using Orbitrap Astral DIA mass spectrometry; (2) establish an atlas of differentially expressed proteins (DEPs) and characterize their functional pathways; and (3) explore the utility by developing a multi-protein diagnostic panel using machine learning approaches.

## 2. Methods

### Study Design and Ethical Approval

This cross-sectional diagnostic accuracy study (Figure 1) was conducted at the Shanghai Ninth People’s Hospital between April 2022 and April 2024. Saliva samples were obtained from a parent diagnostic accuracy trial evaluating point-of-care biomarkers for periodontitis (Li et al., 2025). The study protocol was approved by the Shanghai Ninth People’s Hospital Ethics Committee (IRB Approval: SH9H-2021-T408-3) and was registered at ClinicalTrials.gov (NCT05513599). Written informed consent was obtained from all participants. The study adhered to STARD guidelines.

**FIGURE 1.**
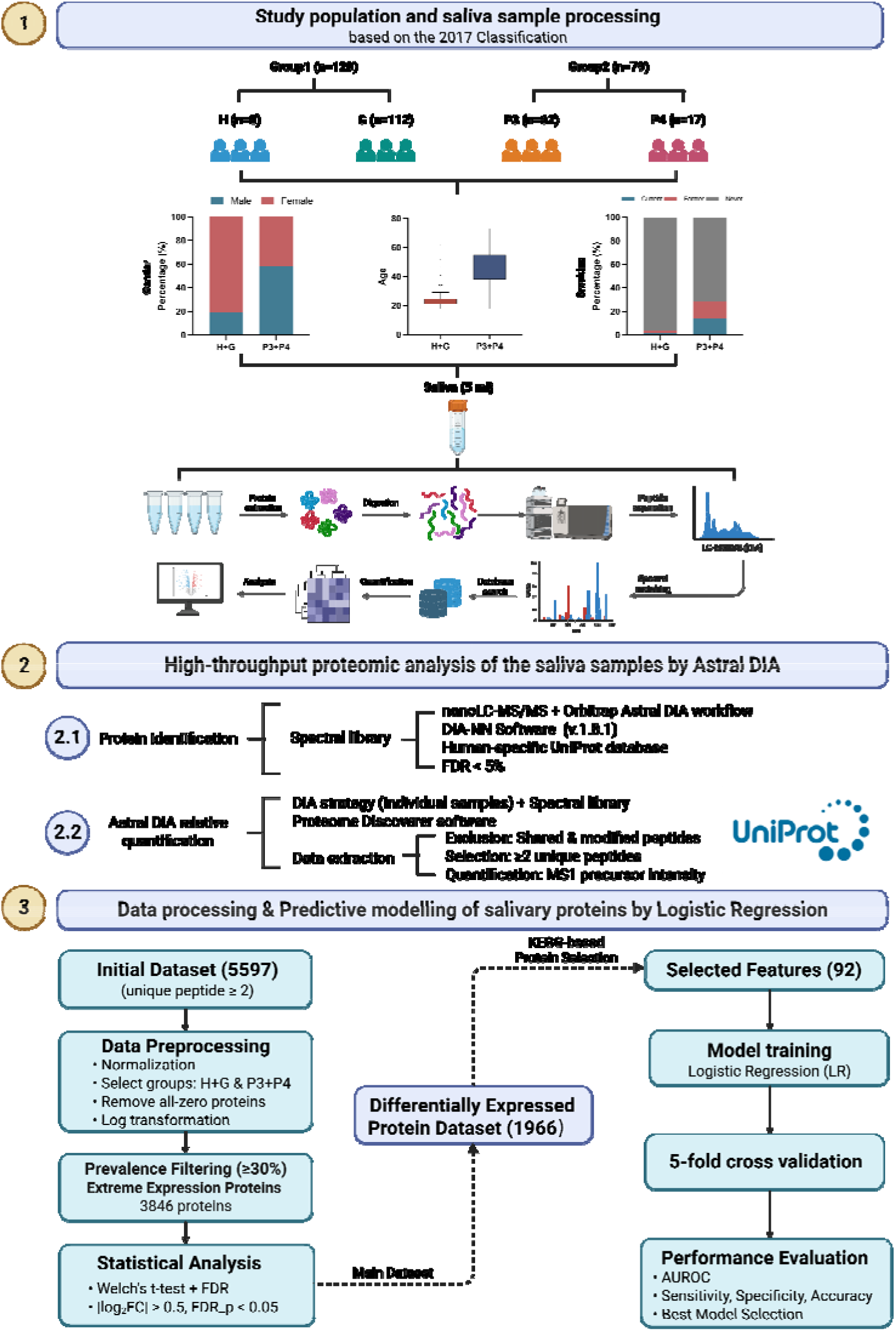
Biomarker Discovery Pipeline. The diagram illustrates the sample collection, saliva processing, high-throughput proteomic analysis, quality control, and data analysis. Group 1 comprises subjects with health (H) and gingivitis (G). P3 and P4 (group 2) are subjects with stage III or IV periodontitis (severe disease). DIA: data-independent acquisition; LC-MS: liquid chromatography-mass spectrometry; FC: fold changes; FDR: false discovery rate; AUROC: Area Under the Receiver Operating Characteristic Curve. See methods for details.

### Study Population and Power

Participants were recruited and classified following previously described criteria (Li et al., 2025). Stage I/II periodontitis cases were excluded to enrich for extreme phenotypes. The final cohort comprised 199 participants: control group (periodontal health/gingivitis, H+G, n = 120) and periodontitis group (stage III or IV periodontitis, P3+P4, n = 79). The study is adequately powered to detect large proteomic effects: assuming a conservative within-group standard deviation of 1.0 on the log2 scale, a log2 fold change of 1.5 (≈2.8-fold) corresponds to an effect size of Cohen’s d ≈1.5, which at a two-sided α=0.05 provides >95% power after false discovery rate control.

### Clinical Examination

Full-mouth periodontal examinations were performed by trained and calibrated specialists following previously described protocols (Li et al., 2025). Staging and grading were determined following the 2017 classification criteria (Tonetti et al., 2018).

### Salivary Samples

Unstimulated whole saliva samples (5 mL) were collected following standardized protocols (Navazesh & Kumar, 2008). Participants fasted and abstained from oral hygiene for ≥2 hours prior to collection. Samples were immediately placed on ice, aliquoted, and stored at −80°C.

### Proteomic Analysis

#### Sample Preparation

Frozen saliva samples were thawed on ice and centrifuged (3,000 rpm, 3 minutes, 4°C) to remove particulate matter. Low-abundance proteins were enriched using magnetic beads (TioMagn™ Max, Evlixir Biotech). Protein concentration was determined using the Bradford assay (Bradford Protein Assay Kit, Biognosys). Proteins were reduced with dithiothreitol (DTT, Sigma-Aldrich), alkylated with iodoacetamide (IAM, Sigma-Aldrich), and digested with trypsin (sequencing grade, Promega) at 37°C for 4 h.

#### Liquid Chromatography–Mass Spectrometry

DIA data were acquired on an Orbitrap Astral mass spectrometer coupled to a Vanquish Neo UHPLC system; detailed acquisition parameters are provided in the Supplementary Methods.

#### Data Processing and Protein Quantification

Raw DIA data were processed using DIA-NN software (version 2.3.2) with a library-free search strategy against the UniProt human proteome database (Demichev, Messner, Vernardis, Lilley, & Ralser, 2020). Peptides and proteins with Q-value < 0.01 were retained.

### Bioinformatics Analysis

#### Differential Expression Analysis

Raw protein intensities were obtained from DIA-NN output, retaining only proteins identified with ≥2 unique peptides. Data were total ion current (TIC) -normalized, followed by log2 transformation. Two categories of DEPs were identified. First, proteins detected in ≥30% of samples within each group were subjected to Welch’s t-test with Benjamini-Hochberg correction; those meeting FDR < 0.05 and |log_2_FC| > 0.5 were considered DEPs. Second, extremely expressed proteins—defined as those detected in ≥70% of samples in one group but ≤30% in the other—were additionally included regardless of statistical testing.

#### Functional Enrichment Analysis

Gene Ontology (GO) and Kyoto Encyclopedia of Genes and Genomes (KEGG) pathway enrichment analyses were performed using the Metascape online platform (Zhou et al., 2019). Enriched terms with adjusted p-values < 0.05 were considered statistically significant. Protein-protein interaction (PPI) networks were constructed using the STRING online database (version 12.0, confidence score ≥ 0.400) and visualized using Cytoscape (version 3.10.3). Hub proteins were identified based on node degree (number of connections).

### Machine Learning-Based Biomarker Selection

#### Feature Selection Strategy

A two-tier feature selection strategy was employed. First, for each of 14 periodontitis-related KEGG pathways, logistic regression identified the top 5 proteins by absolute regression coefficient (Hartman et al., 2023; Nakayasu et al., 2021). Second, individual proteins were evaluated for their diagnostic performance using cross-validated logistic regression, and those achieving an AUC > 0.75 were retained. The combination of proteins from both screening approaches yielded 92 candidate biomarkers for subsequent model development.

#### Model Development and Validation

Logistic regression models were constructed to evaluate the diagnostic performance of multi-protein panels (Shu et al., 2020). For each panel size (2 to 5 proteins), up to 5,000 random protein combinations were sampled from the 92 candidates. Model performance was assessed using 5-fold cross-validation, and the top 10 combinations with the highest mean AUC were selected for each panel size. Final evaluation was performed using a 7:3 train-test split. The optimal panel was selected based on AUC, sensitivity, and specificity.

### Corroboration Using Public Datasets

#### Single-Cell RNA Sequencing Validation

Gene expression patterns were examined using publicly available single-cell RNA sequencing (scRNA-seq) data from human gingival tissues (Easter et al., 2024); dataset details are provided in the Supplementary Methods. Gene expression was visualized and analyzed using the CZ CELLxGENE platform. Differential expression between healthy and periodontitis groups was assessed, with adjusted p-values < 0.05 considered statistically significant.

#### Salivary Proteomics Validation

An independent salivary proteomics dataset was retrieved from the ProteomeXchange Consortium (Deutsch et al., 2025) with the identifier PXD043491 (Blanco-Pintos et al., 2025). This dataset comprised saliva samples from 83 subjects (44 periodontally healthy controls and 39 patients with generalized Stage III-IV periodontitis) analyzed using SWATH-MS. Raw data were reprocessed using DIA-NN (version 1.8.1) with identical parameters. DEPs were identified using the same criteria (|log_2_FC| > 0.5 and FDR < 0.05).

#### Statistical Analysis

Statistical analyses were performed using R (version 4.4.2), and GraphPad Prism (version 10.3.1). Clinical characteristics were compared using Mann-Whitney U test (continuous variables) and Fisher’s exact test (categorical variables). Diagnostic performance of biomarker panels was evaluated using receiver operating characteristic (ROC) curve analysis. All tests were two-tailed, and p-values < 0.05 were considered statistically significant.

## 3. Results

### Study Population Characteristics

Unstimulated whole saliva samples were collected from 199 participants (Table S1). Participants were stratified into two groups: the control group (H+G, n = 120) comprising periodontal health (n = 8) and gingivitis (n = 112), and the periodontitis group (P3+P4, n = 79) comprising Stage III (n = 62) and Stage IV (n = 17) periodontitis. The P3+P4 group was older (46.48 ± 11.79 vs. 23.71 ± 6.06 years), more frequently male (59.5% vs. 19.2%) and more often current/former smokers (27.8% vs. 3.3%) than H+G (all p < 0.001) (Table S1).

### Salivary Proteomic Profiling

Orbitrap Astral DIA-MS analysis quantified a total of 5,597 proteins across all 199 saliva samples. Individual samples yielded an average of 3,588 ± 500 proteins in the H+G group and 3,739 ± 674 proteins in the P3+P4 group, demonstrating comparable proteome depth between groups (Figure S1). Chromatographic stability was confirmed by consistent iRT peptide performance across all runs (Figure S2).

After applying prevalence filtering, 3,846 proteins were retained for differential expression analysis. Comparison between groups identified 1,966 differentially expressed proteins (DEPs), comprising 1,247 upregulated and 719 downregulated proteins in periodontitis (Figure 2A). A subset of DEPs overlapped with reported periodontitis biomarkers (Ahmad, Escalante-Herrera, Marin, & Siqueira, 2025), providing cross-study corroboration (Figure 2A). The PPI network of the top 200 hub proteins ranked by degree among the 1,966 DEPs is shown in Figure 2B. The distribution of DEPs across the log_2_FC spectrum is shown in Figure S3. Principal component analysis (PCA) demonstrated clear separation between groups, with the DEPs subset showing improved discrimination compared to all quantified proteins (Figure S4). The complete list of DEPs is provided in the supplementary Table S2 (“Atlas of periodontitis differentially expressed proteins”).

**FIGURE 2.**
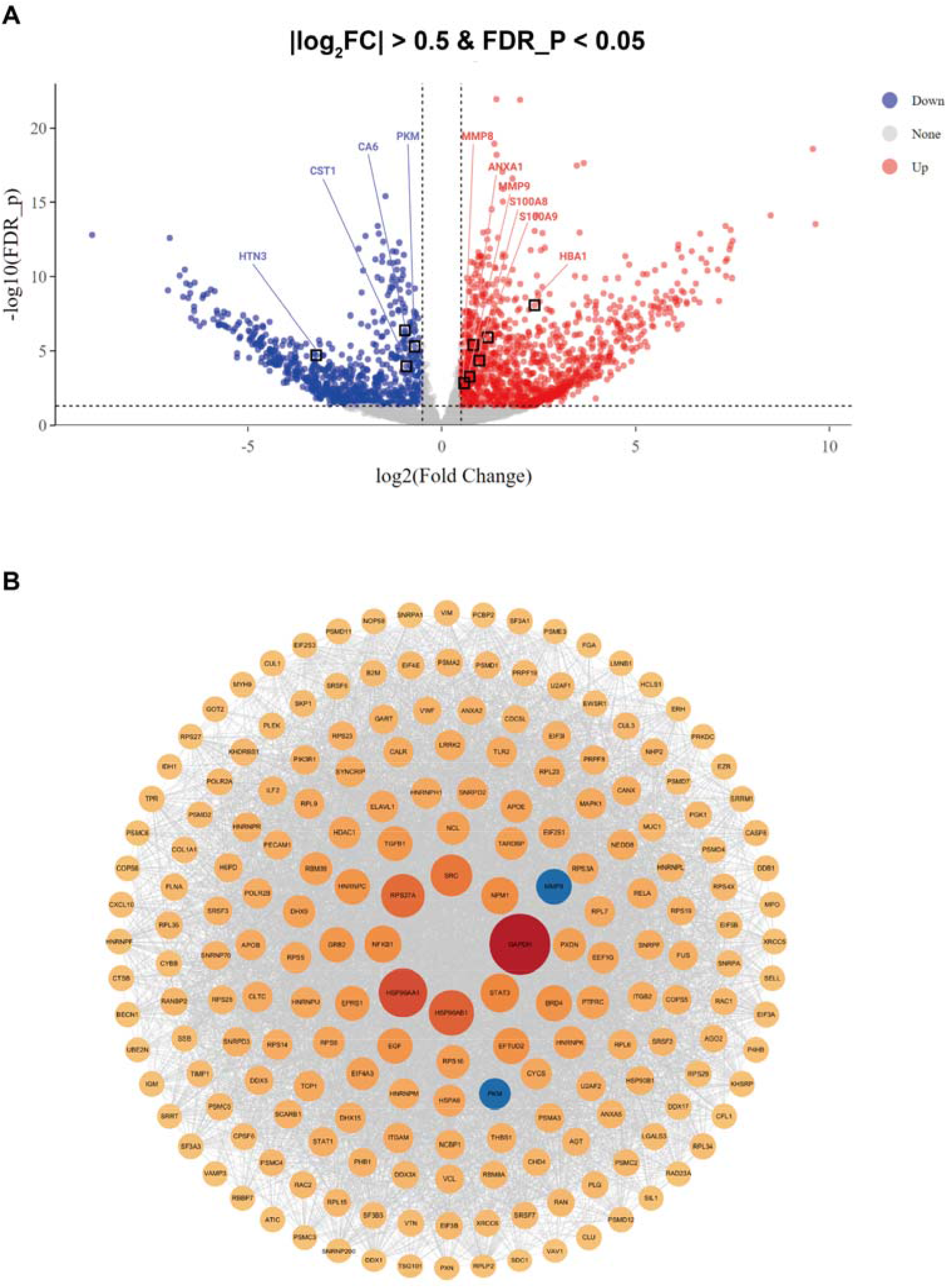
Salivary proteomic profiling of periodontitis. Panel A shows the Volcano plot of differentially expressed proteins between P3+P4 and H+G groups. Red dots indicate proteins upregulated in P3+P4; blue dots indicate proteins downregulated in P3+P4; grey dots indicate non-significant proteins. Black-bordered squares indicate proteins previously reported as periodontitis biomarkers in the literature, identified as differentially expressed in the present dataset, providing cross-study corroboration of our proteomic findings. Panel B illustrated the protein-protein interaction (PPI) network of the top 200 proteins ranked by degree. Degree refers to the number of direct interaction partners a protein has within the network, reflecting its connectivity and functional centrality; node size is proportional to degree. Blue circles indicate proteins previously reported as periodontitis biomarkers in the literature. See also Figure S7 and Table S2. DEPs: differentially expressed proteins. Protein taxonomy is expressed following the NCBI/UniProt guidelines.

### Functional Enrichment Analysis

GO enrichment identified ficolin-1-rich granule, cytoplasmic vesicle lumen, and secretory granule lumen among the top 20 enriched terms (Figure 3A). KEGG analysis identified 48 significantly enriched pathways (Table S3). The top 20 pathways are shown in Figure 3B, with Th17 cell differentiation as the most significant (-log_10_P = 4.10). Among these, 14 periodontitis-relevant pathways were used for subsequent machine learning-based biomarker selection (indicated in Table S3), including Th17 cell differentiation, complement and coagulation cascades, IL-17 signaling pathway, and neutrophil extracellular trap formation. The expression of ficolin-1-rich granule proteins is shown in Figure 3C, with most upregulated in the P3+P4 group. Th17 pathway analysis revealed 15 upregulated and 4 downregulated proteins (Figure 3D), including IKBKG (log_2_FC = 5.18), NFKB1 (log_2_FC = 2.03), STAT3 (log_2_FC = 1.38), TGFB1 (log_2_FC = 1.63), and MAPK14 (log_2_FC = 0.94), while IL1R1 was downregulated (log_2_FC = −4.98).

**FIGURE 3.**
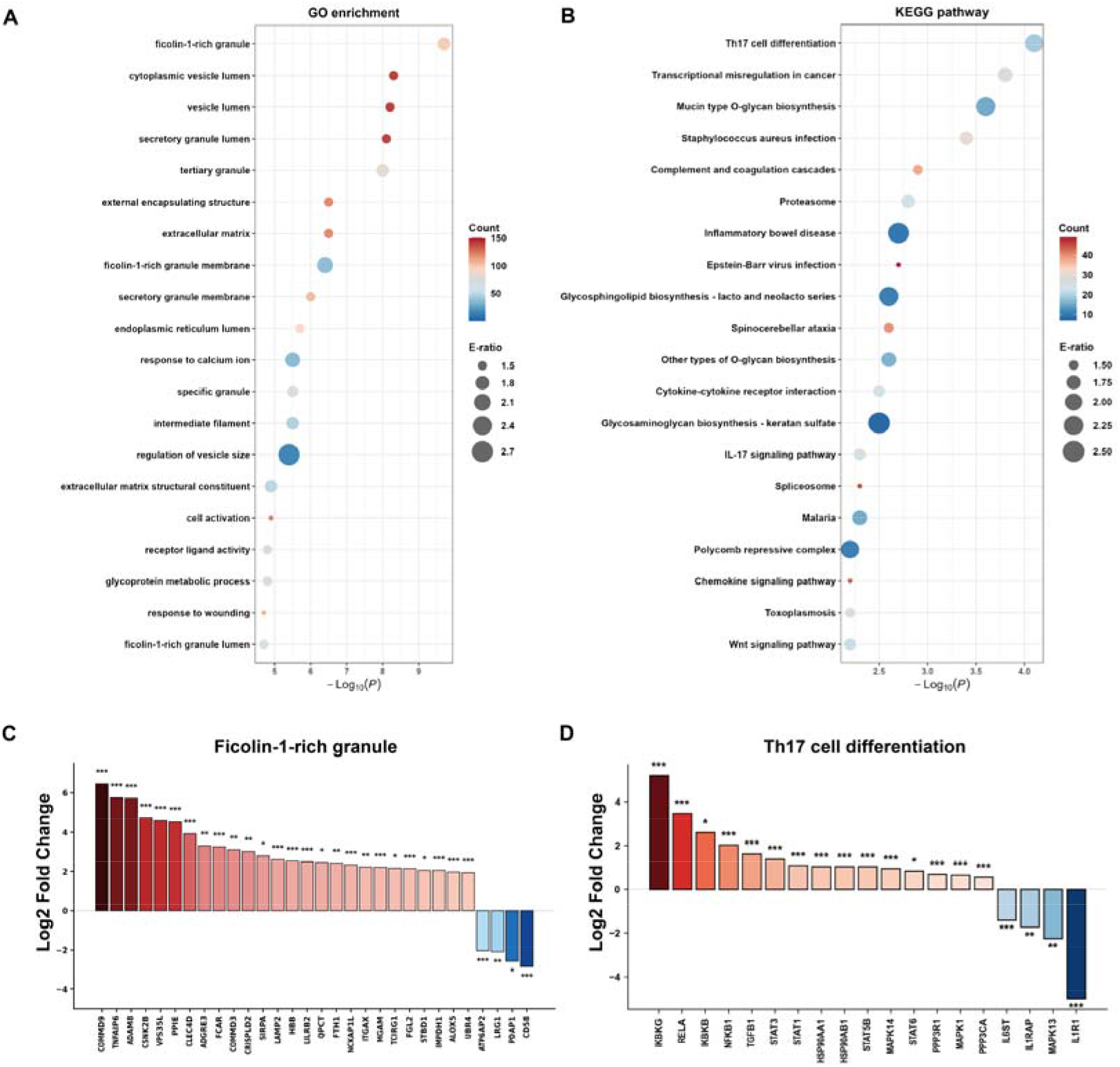
Functional enrichment analysis of differentially expressed proteins. (A) GO enrichment analysis showing the top 20 enriched terms. (B) KEGG pathway enrichment analysis showing the top 20 enriched pathways. Terms are sorted by -Log_10_(P), where P-values were derived from hypergeometric testing with Benjamini-Hochberg correction as implemented in the Metascape platform, and -log10-transformed for visualization, such that higher values indicate greater statistical significance; bubble size indicates E-ratio; color indicates protein count in each pathway. (C) Log_2_FC values of proteins in the ficolin-1-rich granule pathway. (D) Log_2_FC values of proteins in the Th17 cell differentiation pathway. Red: upregulation; blue: downregulation. *FDR < 0.05, **FDR < 0.01, ***FDR < 0.001. FC: fold change; FDR: false discovery rate; GO: Gene Ontology; KEGG: Kyoto Encyclopedia of Genes and Genomes; E-ratio: enrichment ratio, defined as the ratio of observed to expected proportions of proteins annotated by a given term.

### Machine Learning-Based Biomarker Selection

Hierarchical clustering of the 92 candidate biomarkers revealed distinct upregulation and downregulation clusters between groups (Figure 4). The complete heatmap of all 1,966 DEPs is in Figure S5.

**FIGURE 4.**
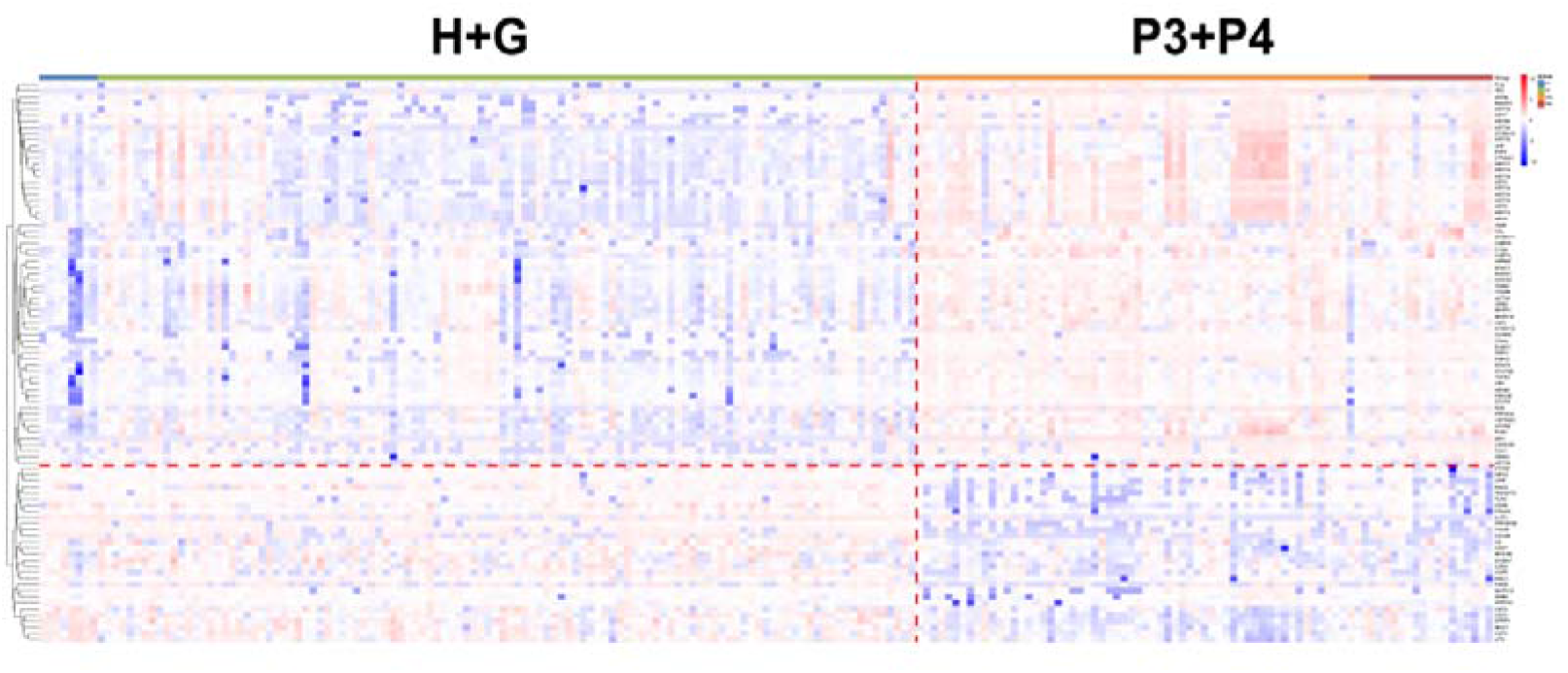
Heatmap of the 92 candidate biomarker proteins between H+G and P3+P4 groups. The heatmap displays normalized protein expression profiles of the 92 candidate biomarker proteins (rows) across all 199 saliva samples (columns), grouped by diagnosis (H+G, left; P3+P4, right). Rows and columns were independently clustered using unsupervised hierarchical clustering with Euclidean distance and complete linkage. Protein expression values were Z-score normalized by row prior to clustering (R package pheatmap). Red indicates higher relative expression; blue indicates lower relative expression. The dashed red line demarcates the two sample groups. H+G: periodontal health and gingivitis; P3+P4: stage III/IV periodontitis.

A two-tier feature selection strategy yielded 92 candidate proteins (Figure 5A). Logistic regression models were constructed to evaluate multi-protein panels ranging from 2 to 5 proteins. Evaluation across panel sizes demonstrated that a 4-protein combination achieved optimal balance between diagnostic performance and model parsimony, with the highest median AUC among all tested configurations (Figure 5B); alternative configurations showed minimal additional performance gain with increased complexity (Figure S6). The best-performing panel comprised TEC (Tec protein tyrosine kinase, upregulated), RAC1 (Rac family small GTPase 1, downregulated), MAPK14 (mitogen-activated protein kinase 14/p38α, upregulated), and KRT17 (keratin 17, upregulated). This four-protein panel achieved an AUC of 0.985 ± 0.010 for distinguishing P3+P4 from H+G groups under 5-fold cross-validation (Figure 5C). Confusion matrix analysis of the independent test set showed 100% specificity for the H+G group and 83% sensitivity for the P3+P4 group (Figure 5D). Their distribution across the proteome is shown in Figure 5E.

**FIGURE 5.**
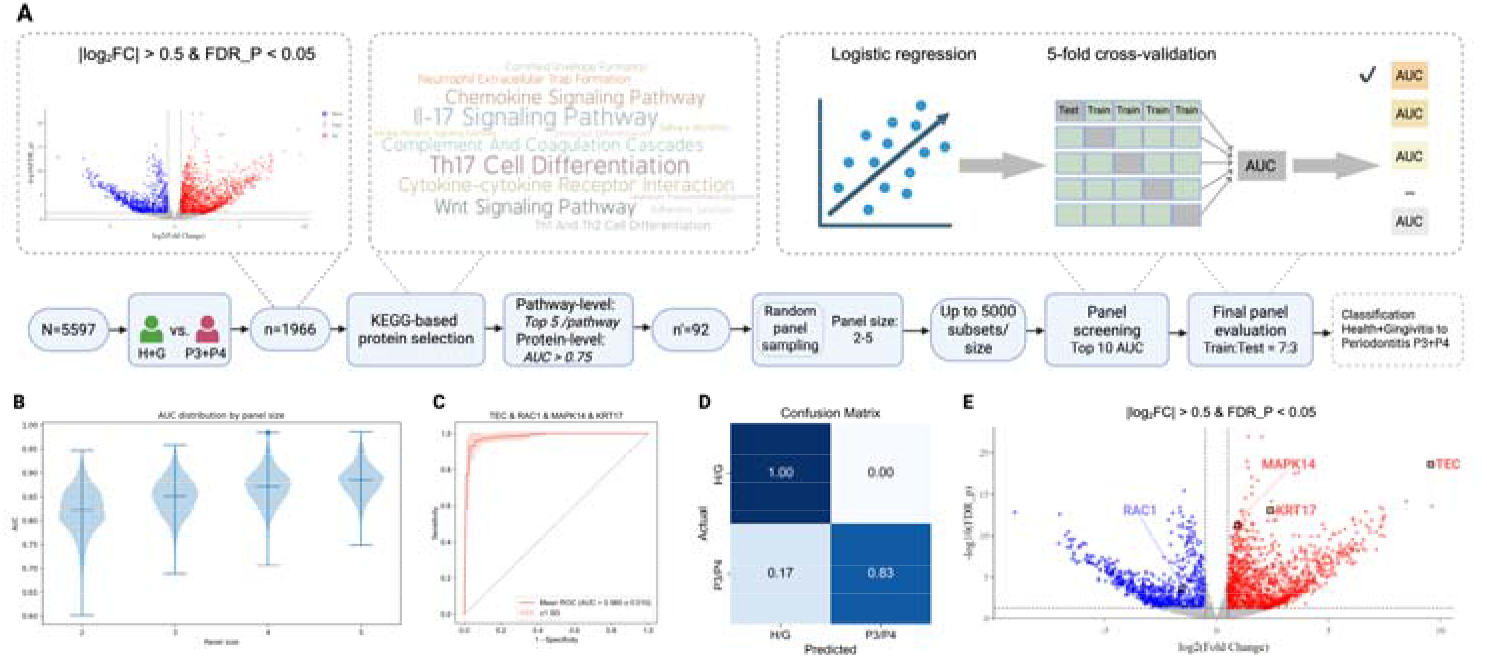
Identification of optimal biomarker panel using machine learning strategy. (A) The workflow of biomarker selection, including pathway-level screening and individual protein AUC evaluation to prioritize candidate biomarkers. (B) Violin plot comparing diagnostic performance (AUC) of multi-protein panels with different sizes (2 to 5 proteins). (C) ROC curves of the four-protein panel — TEC (Tec protein tyrosine kinase), RAC1 (Rac family small GTPase 1), MAPK14 (mitogen-activated protein kinase 14), and KRT17 (keratin 17) — from 5-fold cross-validation. (D) Confusion matrix of the four-protein panel on the independent test set, demonstrating 100% specificity for the H+G group and 83% sensitivity for the P3+P4 group. (E) Volcano plot of differentially expressed proteins between P3+P4 and H+G groups, with the four proteins of the optimal diagnostic panel highlighted. Red dots indicate upregulated proteins; blue dots indicate downregulated proteins in P3+P4. Black-bordered squares indicate TEC (upregulated), RAC1 (downregulated), MAPK14 (upregulated), and KRT17 (upregulated); full protein annotations are available in the UniProt database (https://www.uniprot.org). See also Figure S6. AUC: area under the curve; ROC: receiver operating characteristic.

### Protein-Protein Interaction Network Analysis

PPI network analysis revealed highly significant enrichment of interactions (p < 1.0×10^−16^) (Figure 6A). The network was visualized using a degree-sorted concentric circle layout in Cytoscape (Figure 6B), with hub proteins (degree ≥ 20) in the innermost circle comprising STAT3, TGFB1, NFKB1, STAT1, MAPK1, RAC2, ITGAM, KRT5, GRB2, and TLR2. Among the four-protein diagnostic panel, MAPK14 (degree=19) and RAC1 (degree=15) occupied the second circle, KRT17 (degree=13) the third circle, and TEC (degree=1) the outermost circle. The complete network is in Figure S7.

**FIGURE 6.**
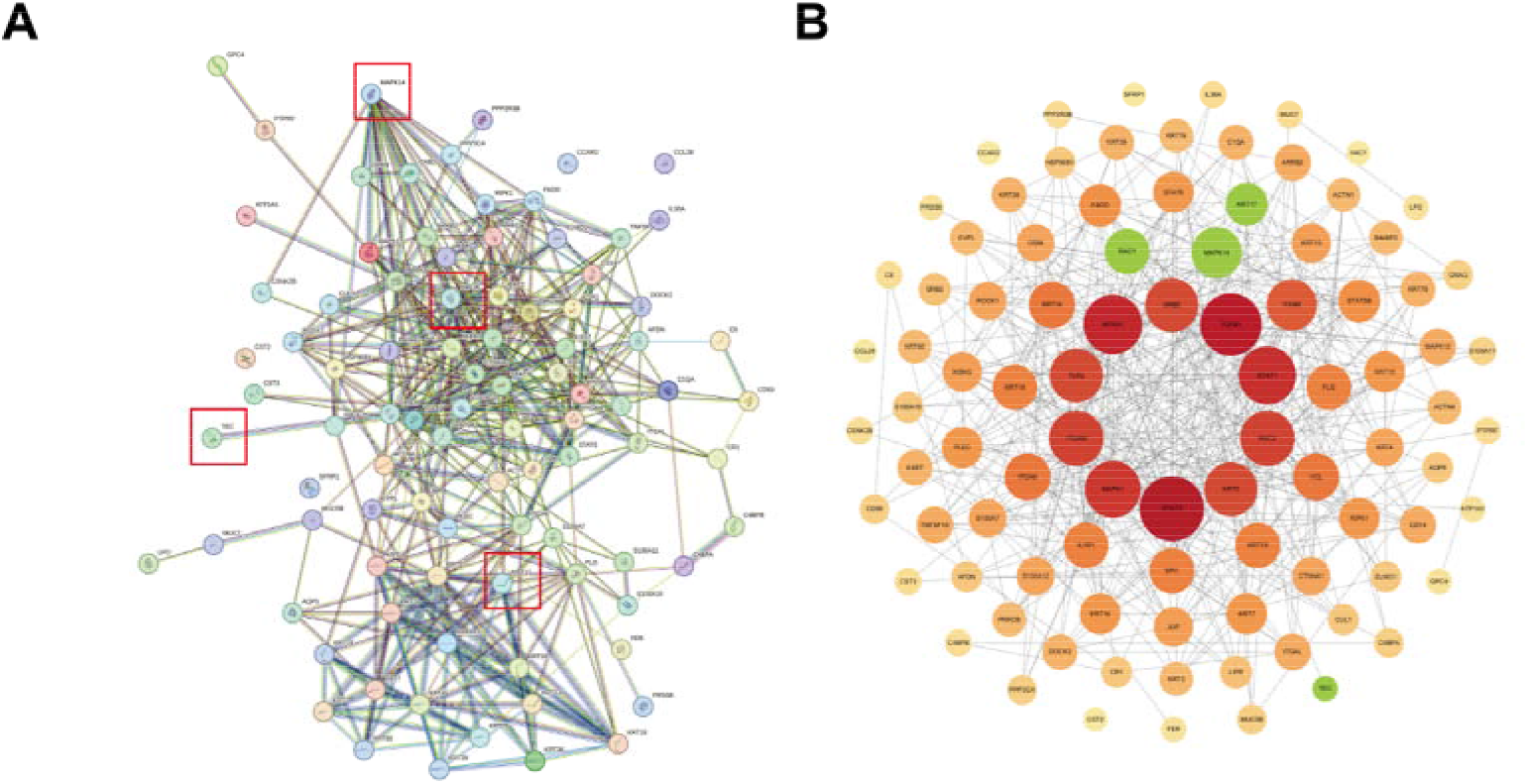
Protein-protein interaction network analysis of the 92 candidate biomarkers. (A) PPI network of the 92 candidate biomarkers constructed using the STRING database (confidence score ≥ 0.400). Each node represents a protein; node colors are assigned by STRING to distinguish individual proteins. Edges represent protein-protein associations, with line colors indicating the type of supporting evidence: cyan, curated databases; pink, experimentally determined; green, gene neighborhood; red, gene fusion; dark blue, gene co-occurrence; yellow-green, text mining; black, co-expression; light blue, protein homology. Red-bordered squares indicate the four proteins of the diagnostic panel (TEC, RAC1, MAPK14, KRT17). (B) Degree-sorted concentric circle layout visualization generated using Cytoscape. Proteins are arranged by degree — defined as the number of direct interaction partners within the network — with hub proteins of highest degree positioned in the innermost circle. Node color reflects connectivity degree, ranging from yellow (low) to red (high). The four biomarkers in the diagnostic panel are highlighted in green. PPI: protein-protein interaction.

### External Validation with Single-Cell RNA Sequencing

Expression patterns of TEC, RAC1, KRT17, and MAPK14 were examined across cell types in H+G and P3+P4 gingival tissues using publicly available scRNA-seq data (Easter et al., 2024) (Figure 7). TEC showed the most pronounced differential expression in mast cells. RAC1 and MAPK14 exhibited broad expression with cell-type-specific mean expression shown for neutrophils and gamma-delta T cells, respectively; KRT17 was predominantly detected in keratinocyte clusters with higher expression in the P3+P4 group (Figure 7). At the global tissue level, TEC was significantly upregulated (log_2_FC = 1.114, adjusted p = 0.015) and RAC1 was significantly downregulated (log_2_FC = −0.455, adjusted p = 0.0005), while KRT17 and MAPK14 did not reach statistical significance (adjusted p = 0.655 and 0.536, respectively) (Table S4).

**FIGURE 7.**
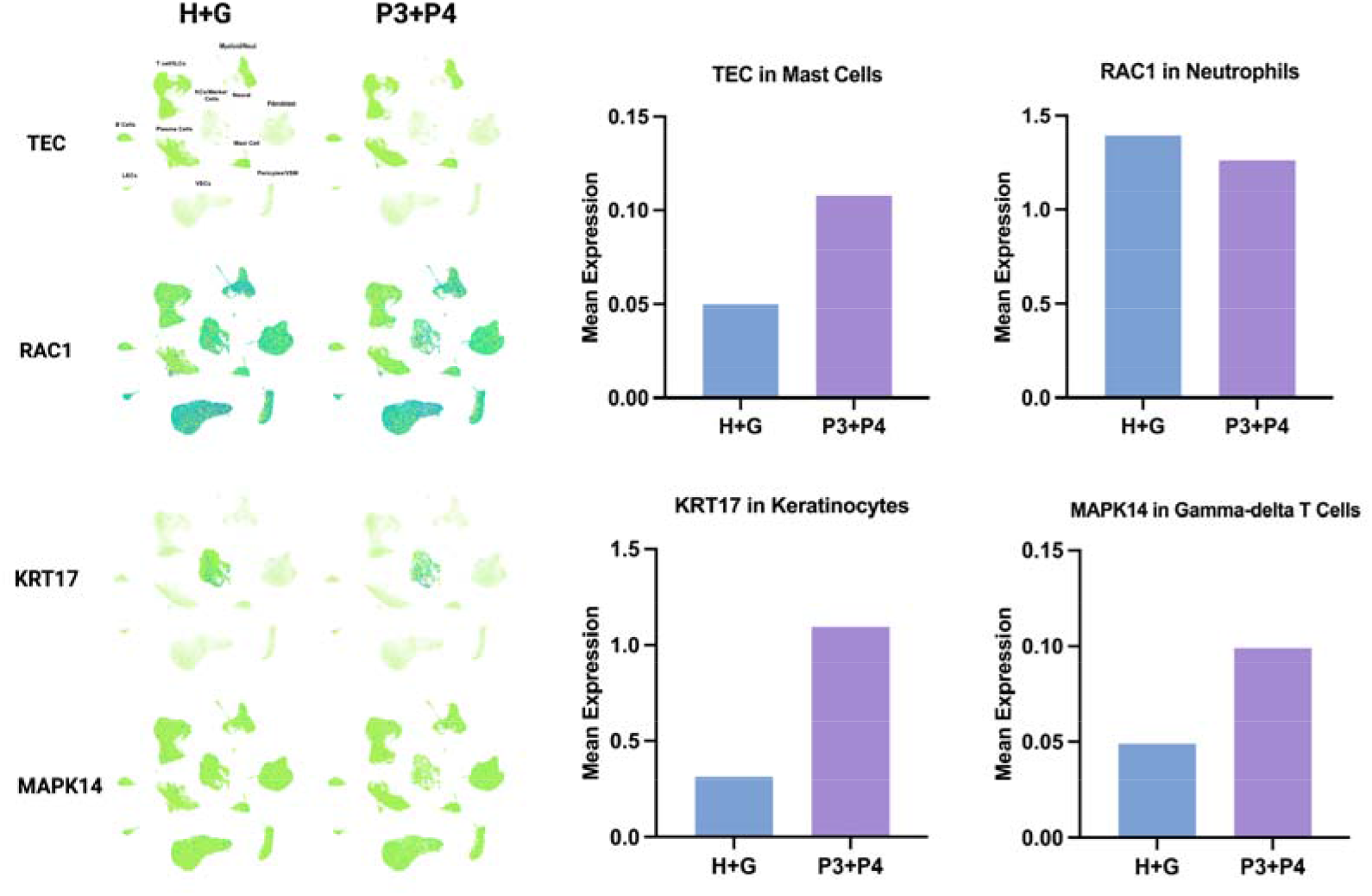
External validation of the four-protein biomarker panel using single-cell RNA sequencing data. (Left) UMAP visualization of TEC, RAC1, KRT17, and MAPK14 gene expression across single cells from H+G and P3+P4 groups, derived from a publicly available human gingival tissue scRNA-seq atlas (GEO: GSE152042, GSE161267, GSE164241, GSE266897) and visualized using the CZ CELLxGENE platform. Each dot represents a single cell; cell color indicates normalized gene expression level on a continuous scale from green (low or absent expression) to blue-purple (high expression). (Right) Bar charts showing mean expression of each biomarker in a representative cell type between H+G and P3+P4 groups: TEC in mast cells, RAC1 in neutrophils, KRT17 in keratinocytes, and MAPK14 in gamma-delta T cells. Mean expression is defined as the total log-normalized gene expression within a cell cluster divided by the total number of cells in that cluster. See also Table S4. scRNA-seq: single-cell RNA sequencing; UMAP: Uniform Manifold Approximation and Projection.

### External Validation Using ProteomeXchange Dataset

Using the independent salivary proteomic dataset (PXD043491), 274 DEPs were identified, including 104 upregulated and 170 downregulated proteins (Table S5). Among the four-protein biomarker panel, TEC, RAC1, and MAPK14 were not detected in this dataset, likely reflecting differences in proteome depth between the Orbitrap Astral and SWATH-MS platforms; KRT17 was detected and showed significant differential expression consistent with our primary findings (log_2_FC = 3.50, FDR = 1.04 × 10^−6^). KEGG pathway analysis of the 274 DEPs revealed enrichment in 14 pathways (Table S6), of which 4 overlapped with our primary analysis: IL-17 signaling pathway, cornified envelope formation, and two infection-related pathways (Staphylococcus aureus infection and tuberculosis) that share downstream innate immune and barrier defense components with periodontitis pathogenesis.

PPI network analysis revealed significant functional connectivity (Figure S8) with MAPK14 (19 interacting DEPs), RAC1 (17 DEPs), and KRT17 (16 DEPs) demonstrating extensive connections, with RAC1-IQGAP1 and KRT17-SFN showing highest confidence (>0.9); TEC exhibited limited connectivity (1 DEP) (Table S7).

## 4. Discussion

This study represents the first application of Orbitrap Astral DIA mass spectrometry for comprehensive salivary proteome profiling in periodontitis. We quantified 5,597 proteins across 199 saliva samples and identified 1,966 differentially expressed proteins between periodontal health/gingivitis and stage III/IV periodontitis, establishing a periodontitis salivary proteomic atlas. A two-tier machine learning strategy identified a four-protein biomarker panel (TEC, RAC1, MAPK14, and KRT17) that achieved high diagnostic accuracy (AUC = 0.985±0.010) for distinguishing stage III/IV periodontitis from periodontal health and gingivitis. Single-cell RNA analyses and available proteomic data provided external corroboration of the findings.

This study offers three methodological advances over prior salivary proteomics investigations. First, the Orbitrap Astral DIA-MS platform enabled quantification of 5,597 proteins, substantially exceeding previous studies (Blanco-Pintos et al., 2025; Grant et al., 2022; Hartenbach et al., 2020; Romano et al., 2023): Blanco-Pintos et al. quantified 377 proteins using SWATH-MS (Blanco-Pintos et al., 2025). Second, individual-level proteomic quantification of 199 participants exceeds most previous investigations, which typically used pooled samples or cohorts of <50 subjects (Grant et al., 2022; Hartenbach et al., 2020; Salazar et al., 2013; Wu, Shu, Luo, Ge, & Xie, 2009). Third, the two-tier machine learning strategy—pathway-based filtering followed by cross-validation—improves upon differential expression analysis.

The four-protein panel achieved an AUC of 0.985 ± 0.010 with 83% sensitivity and 100% specificity. Single biomarkers show limited utility; for instance, aMMP-8 point-of-care tests achieved pooled sensitivity of only 0.59 (Li et al., 2025), and S100A8 achieved an AUC of 0.71 (Corana et al., 2024). Multi-protein panels have shown improved performance: Bostanci et al.’s five-protein panel (MMP9, RAP1A, ARPC5, CLUS, DMBT1) reached AUC of 0.97 (Bostanci et al., 2018), Grant et al.’s four-protein panel (MMP9, S100A8, A1AGP, PK) achieved AUC of 0.96 (Grant et al., 2022), and Blanco-Pintos et al.’s two-protein combination (B2M, PFN1) achieved 96.3% accuracy (Blanco-Pintos et al., 2025). Our panel achieved comparable accuracy while utilizing novel biomarker combinations not previously reported in systematic reviews (Ahmad et al., 2025; Corana et al., 2024; Hu & Leung, 2023), reflecting the expanded proteome coverage enabled by nanoparticle-bead enrichment and Orbitrap Astral DIA-MS. Notably, seven of these ten unique biomarkers (MMP9, RAP1A, CLUS, S100A8, A1AGP, PK, B2M) were also identified as DEPs in our dataset, providing independent cross-study validation of our proteomic findings.

Biologically, the four biomarkers represent distinct yet mechanistically interconnected nodes within a self-amplifying inflammatory cascade in periodontitis pathogenesis, providing a biological rationale for the panel’s diagnostic robustness. At the upstream level, RAC1 and TEC modulate innate immune defense and bone resorption, respectively. RAC1, functioning as a key regulator of the MEKK3-MKK3/6-p38 MAPK cascade, is essential for neutrophil antimicrobial functions, including extracellular trap formation (Gavillet, Martinod, Renella, Wagner, & Williams, 2018; Uhlik et al., 2003). Its significant downregulation—confirmed in both our salivary proteome and the scRNA-seq atlas—is consistent with murine evidence linking Rac-deficient leukocytes to impaired pathogen clearance and accelerated alveolar bone loss (Sima, Gastfreund, Sun, & Glogauer, 2014). TEC, showing the most pronounced upregulation in our dataset, drives osteoclastogenesis through RANKL-RANK signaling and PLCγ-mediated calcium signaling (Shinohara et al., 2008), directly linking it to the hallmark bone destruction of stage III/IV periodontitis. At the single-cell level, the most pronounced differential TEC expression was observed in mast cells (Huang, Lu, Chen, Huang, & Liu, 2013), which are detected in higher density in periodontal disease, consistent with the established role of TEC-family kinases in pro-inflammatory mediator release in periodontal tissues, and further supported by its significant transcriptional upregulation in the scRNA-seq atlas (log_2_FC = 1.114, adjusted p = 0.015).

These upstream signals converge on MAPK14 (p38α), which serves as a central inflammatory hub. In our dataset, MAPK14 was enriched across 14 periodontitis-relevant pathways and formed an extensive signaling network with key transcription factors—including NFKB1, NFKB2, RELA, STAT3, STAT1, and TGFB1—all significantly upregulated, consistent with its established role as a convergence point of inflammatory signaling in periodontal disease (Kirkwood & Rossa, 2009; Travan, Li, D’Silva, Slate, & Kirkwood, 2013). Notably, both RAC1 (via MEKK3-MKK3/6) and TEC (via PLCγ-calcium signaling) can activate p38 MAPK, positioning MAPK14 as an integration point of the upstream signals.

Downstream of this inflammatory convergence, KRT17 reflects the resulting epithelial barrier compromise. Regulated by the NFκB/MAPK and STAT1/3 pathways activated by MAPK14 (Romashin et al., 2024), KRT17 functions as a “barrier alarmin” (Zhang, Yin, & Zhang, 2019) and participates in a bidirectional regulatory loop with STAT3 (Yang et al., 2018). Its upregulation is consistent with spatially resolved transcriptomic data demonstrating broader KRT17 expression across junctional keratinocytes in diseased periodontal tissues (Easter et al., 2024), and it was the only panel biomarker detected and confirmed in the independent SWATH-MS dataset, providing cross-platform proteomic corroboration.

Collectively, this mechanistic architecture—neutrophil dysregulation (RAC1), bone resorption (TEC), inflammatory signal convergence (MAPK14), and epithelial barrier compromise (KRT17)—captures the multidimensional pathogenesis of advanced periodontitis, consistent with pathway meta-analyses (Ahmad et al., 2025) and likely underlies the panel’s diagnostic robustness.

The diagnostic potential of salivary proteins is grounded in the compositional relationship between saliva and gingival crevicular fluid (GCF) (Buzalaf et al., 2020; Giannobile et al., 2009). Recent proteomic mapping demonstrated 685 proteins shared between the two biofluids, with sample type accounting for only 2.8% of proteome variation (Bao et al., 2025). Notably, MAPK14 and KRT17 were detected in GCF proteomics, with KRT17 showing high abundance in both biofluids (Bao et al., 2025), supporting their translational relevance. Unlike single-biomarker approaches such as aMMP-8, which demonstrate limited sensitivity (Bi et al., 2025; Deng et al., 2022), this multi-protein panel captures distinct pathogenic dimensions, potentially improving diagnostic robustness, and could be translated into point-of-care devices for chairside use or population screening.

Several limitations warrant consideration. The extreme phenotype design—comparing health/gingivitis with stage III/IV periodontitis—maximizes discriminatory power but may inflate diagnostic performance estimates and leaves the panel’s capacity to detect early-stage disease undetermined. External validation was also limited in scope: three of the four panel proteins (TEC, RAC1, MAPK14) were undetectable in the independent SWATH-MS dataset due to platform-dependent proteome depth differences, and only TEC and RAC1 reached transcriptional significance in the scRNA-seq atlas.

Future research should prioritize: validation across the full disease severity spectrum; immunoassay validation (e.g., ELISA) to confirm protein-level detectability; and replication using comparable DIA-MS platforms to resolve platform-dependent detection of TEC, RAC1, and MAPK14.

## Conclusion

This study presents the first comprehensive salivary proteomic atlas of periodontitis, generated using Orbitrap Astral DIA mass spectrometry, and identifies 1,966 DEPs. A four-protein diagnostic panel comprising TEC, RAC1, MAPK14, and KRT17—representing neutrophil dysregulation, bone resorption, inflammatory signal convergence, and epithelial barrier compromise—achieved an AUC of 0.985 with 83% sensitivity and 100% specificity. External corroboration from single-cell transcriptomic and independent proteomic datasets supports the biological plausibility of the panel.

## Data Availability

Some of the data are available online, other data are not available due to confidentiality issues

https://cellxgene.cziscience.com/collections/71f4bccf-53d4-4c12-9e80-e73bfb89e398

## Author contributions

**Xiaoyu Yu** and **Runting Yan** contributed to study design, data analysis and interpretation, and led manuscript preparation. **Hairui Li, Yu Xie, Mengning Bi, Yuan Li, Andrea Roccuzzo** contributed to data analysis and interpretation. **Maurizio S. Tonetti** devised this study and contributed to protocol development, data analysis, data interpretation, and manuscript preparation. All authors critically revised the manuscript, gave their final approval, and agreed to be accountable for all aspects of the work.

## Acknowledgements

The authors thank all participants for their contribution to this study, and the clinical staff of Shanghai Ninth People’s Hospital for their assistance with sample collection.

## Data Availability Statement

The mass spectrometry proteomics data have been deposited to the Proteom eXchange Consortium via the PRIDE partner repository with the dataset identi fier PXD072303. The integrated single-cell RNA sequencing atlas used for valid ation is publicly available at CZ CELLxGENE (https://cellxgene.cziscience.com/collections/71f4bccf-53d4-4c12-9e80-e73bfb89e398), with original raw data from t he four integrated studies deposited in GEO (GSE152042, GSE161267, GSE1642 41, GSE266897). The external salivary proteomics dataset used for validation i s available in ProteomeXchange under identifier PXD043491.

