## Supplementary Material for "Severe Periodontitis Biomarker Identification by Deep Salivary Proteome Profiling with Astral DIA Mass Spectrometry"

**TABLE S1 Demographics, periodontal parameters, and medical co-morbidities in the whole population and by different periodontal diagnoses.**

|  | |  | **All**  **(n=379)** | **Health/**  **Gingivitis (n=120)** | **Stage I/II Periodontitis (n=180)** | **Stage III/IV Periodontitis (n=79)** | ***p* value** |
| --- | --- | --- | --- | --- | --- | --- | --- |
| **Age** | |  | 29.81 ± 12.03 | 23.71 ± 6.06 | 26.55 ± 7.95 | 46.48 ± 11.79 | <0.001 |
| **Sex** | | | | | |  | <0.001 |
|  | Male | | 130 (34.3%) | 23 (19.2%) | 60 (33.3%) | 47 (59.5%) |  |
|  | Female | | 249 (65.7%) | 97 (80.8%) | 120 (66.7%) | 32 (40.5%) |  |
| **Education level** | | | | | |  | <0.001 |
|  | No Education/Elementary School/Junior High School | | 10 (2.6%) | 0 (0.0%) | 3 (1.7%) | 7 (8.9%) |  |
|  | Senior high school  /Technical secondary school | | 19 (5.0%) | 3 (2.5%) | 10 (5.6%) | 6 (7.6%) |  |
|  | Junior college/Bachelor | | 231 (60.9%) | 65 (54.2%) | 109 (60.6%) | 57 (72.2%) |  |
|  | Master/Doctor | | 119 (31.4%) | 52 (43.3%) | 58 (32.2%) | 9 (11.4%) |  |
| **Family income ^a^** | | | | | |  | 0.399 |
|  | Low income | | 83 (21.9%) | 27 (22.5%) | 44 (22.4%) | 12 (15.2%) |  |
|  | Medium income | | 197 (52.0%) | 57 (47.5%) | 97 (53.9%) | 43 (54.4%) |  |
|  | High income | | 64 (16.9%) | 22 (18.3%) | 24 (13.3%) | 18 (22.8%) |  |
|  | Don't know | | 35 (9.2%) | 14 (11.7%) | 15 (8.3%) | 6 (7.6%) |  |
| **Diabetes status** | | | | | |  | 0.156 |
|  | No | | 377 (99.5%) | 120 (100.0%) | 180 (100.0%) | 77 (97.4%) |  |
|  | Yes | | 2 (0.5%) | 0 (0.0%) | 0 (0.0%) | 2 (2.6%) |  |
| **Smoking status** | | | | | |  | <0.001 |
|  | Never | | 339 (89.4%) | 116 (96.7%) | 166 (92.2%) | 57 (72.2%) |  |
|  | Former smoker | | 17 (4.5%) | 2 (1.7%) | 4 (2.2%) | 11 (13.9%) |  |
|  | Current smoker | | 23 (6.1%) | 2 (1.7%) | 10 (5.6%) | 11 (13.9%) |  |
|  | Heavy smoker ^b^ | | 14 (3.7%) | 0 (0.0%) | 4 (2.2%) | 10 (12.7%) |  |
| **Alcohol consumption** | | | | | |  | 0.006 |
|  | No | | 153 (40.4%) | 54 (45.0%) | 77 (42.8%) | 22 (27.8%) |  |
|  | Yes | | 222 (58.6%) | 66 (55.0%) | 102 (56.7%) | 54 (68.4%) |  |
|  | Quitted | | 4 (1.1%) | 0 (0.0%) | 1 (0.6%) | 3 (3.8%) |  |
| **Clinical parameters** | | | | | |  |  |
|  | PPD (mean ± SD) | | 3.09 ± 1.05 | 2.88 ± 0.81 | 3.07 ± 0.92 | 3.47 ± 1.49 | <0.001 |
|  | CAL (mean ± SD) | | 0.70 ± 1.53 | 0.05 ± 0.30 | 0.25 ± 0.58 | 2.88 ± 2.25 | <0.001 |
|  | Furcation involved teeth (class II & III) per patient (mean ± SD) | | 0.54 ± 1.37 | 0.00 ± 0.00 | 0.00 ± 0.00 | 2.56 ± 1.95 | <0.001 |
|  | Mobile teeth  (class II & III) per patient (mean ± SD) | | 0.17 ± 1.11 | 0.00 ± 0.00 | 0.00 ± 0.00 | 0.82 ± 2.33 | <0.001 |
|  | BOP percentage per patient (mean ± SD) | | 59.12 ± 18.91 | 49.48 ± 18.50 | 61.70 ± 17.13 | 67.90 ± 17.36 | <0.001 |

*Notes*:

^a^ For family income, low income means family income of RMB 0-5000/person per month, medium income means family income of RMB 5000-15000/person per month, high income means family income of RMB >15000/person per month.

^b^ Heavy smoker means smoking more than 10 cigarettes per day.

P values represent comparisons between Health/Gingivitis group and Stage III/IV Periodontitis group.

**TABLE S2 Atlas of periodontitis differentially expressed proteins**

The list is provided in Microsoft Excel format.

**TABLE S3** **KEGG pathway enrichment analysis results for the 1,966 differentially expressed proteins.**

The list is provided in Microsoft Excel format.

**TABLE S4 Validation of machine learning-identified proteins using single-cell RNA sequencing data from periodontitis atlas.**

| **Gene** | **Periodontitis vs Healthy** | |
| --- | --- | --- |
|  | **log_2_FC** | **P_value adjust** |
| **TEC** | 1.114 | 0.01494 |
| **KRT17** | 0.393 | 0.655 |
| **MAPK14** | 0.06869 | 0.5355 |
| **RAC1** | -0.455 | 0.0004962 |

*Notes*:

The log_2_FC and p-values were obtained from Easter et al. (2024) Supplementary Data 1, which compared periodontitis versus healthy samples.

**TABLE S5 Differentially expressed proteins identified from the external salivary proteomics validation dataset (ProteomeXchange: PXD043491).**

The list is provided in Microsoft Excel format.

**TABLE S6 KEGG pathway enrichment analysis results for the 274 DEPs in the external salivary proteomics validation dataset (ProteomeXchange: PXD043491).**

The list is provided in Microsoft Excel format.

**TABLE S7 Protein-protein interactions between the four-protein biomarker panel and DEPs from the external validation dataset.**

| **Protein** | **Interacting DEPs (n)** | **High-confidence interactions (combined score)** |
| --- | --- | --- |
| **MAPK14** | 19 | RAC1, HSP90AB1, GSTP1 (medium to high confidence, 0.6-0.7) |
| **RAC1** | 17 | IQGAP1(highest confidence, >0.9)  ARHGDIB, ARHGAP1, ACTB, MYL6, MYL9 (high confidence, >0.7) |
| **KRT17** | 16 | SFN (highest, >0.9)  KRT5, KRT6A, KRT6B, KRT16, KRT14 (high-highest confidence, 0.8-0.9) |
| **TEC** | 1 | IGKVD-28 only (medium to high confidence, 0.5) |

*Notes:*

Combined score represents the confidence of protein-protein interactions from STRING database, ranging from 0 to 1.

Scores are classified as: highest confidence (> 0.9), high confidence (> 0.7), and medium confidence (> 0.4).

**FIGURE S1 Distribution of quantified unique peptides and proteins per sample in the H+G and P3+P4 groups.**

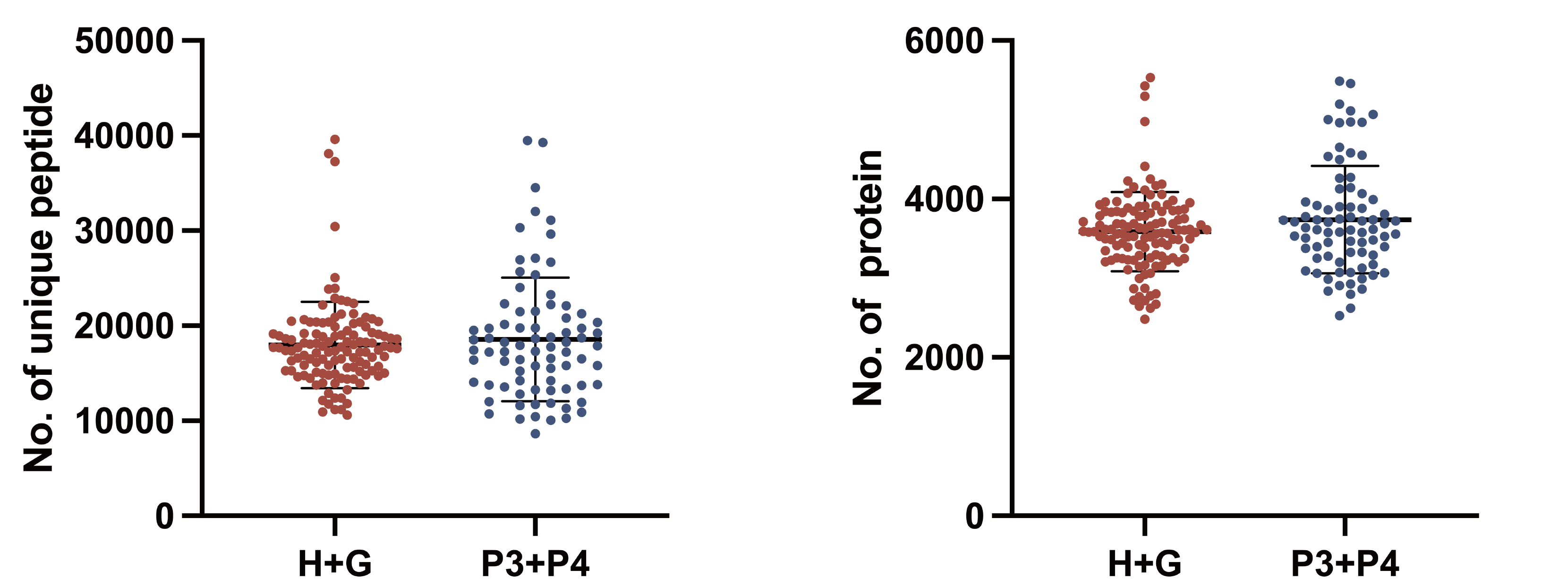

Distribution of quantified unique peptides and proteins per sample in the H+G and P3+P4 groups. Each dot represents one individual saliva sample. Lines indicate the median and interquartile range. H+G: n=120; P3+P4: n=79.

**FIGURE S2 Quality control assessment of mass spectrometry analysis.**

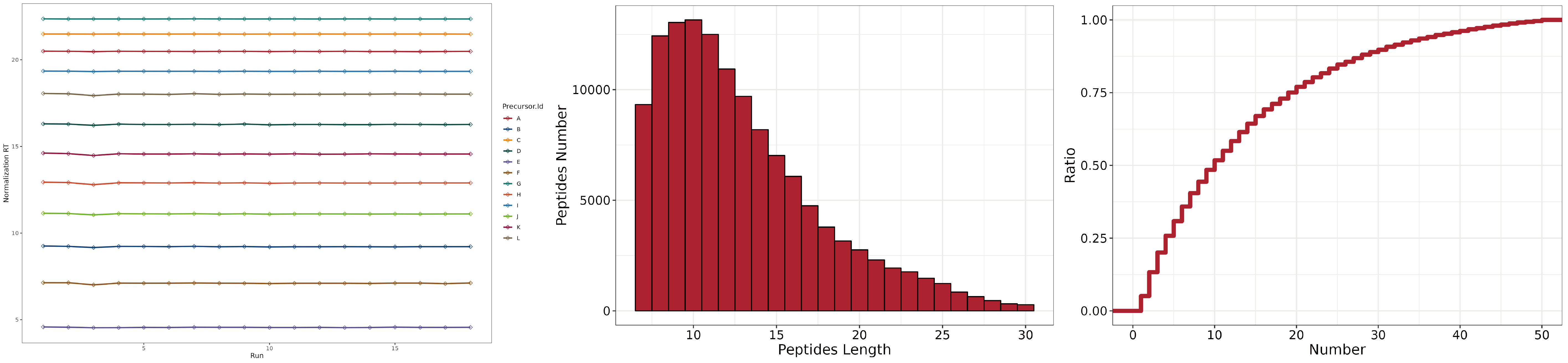
 (Left) Indexed retention time (iRT) peptide performance across all sample runs; each line represents one iRT reference peptide, demonstrating consistent retention time alignment across runs. (Middle) Distribution of identified peptide lengths across all samples. (Right) Cumulative distribution of identified peptide counts per sample.

**FIGURE S3 Distribution of differentially expressed proteins (DEPs) across the |log_2_FC| spectrum.**

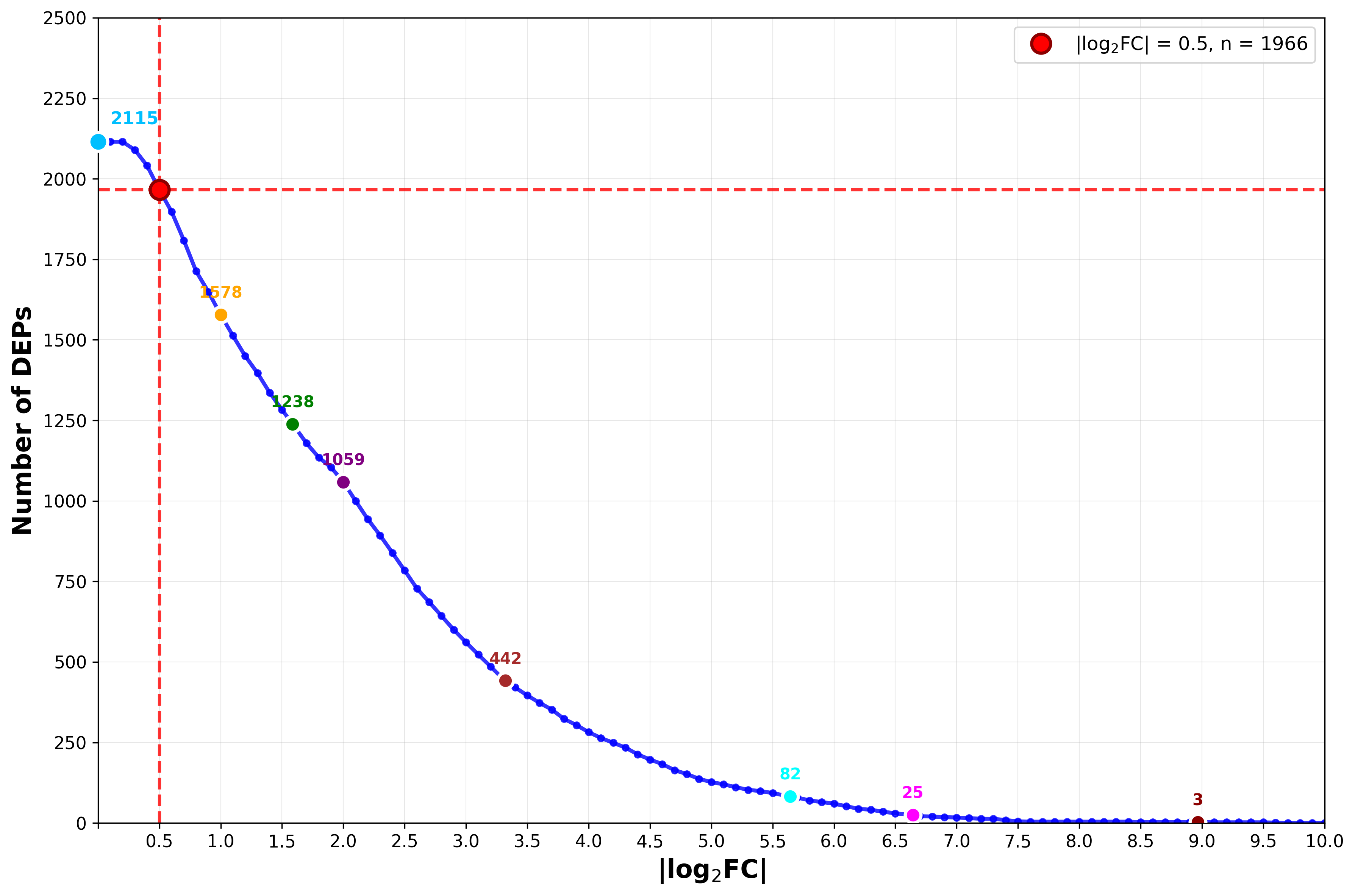

The x-axis represents the |log_2_FC| threshold and the y-axis indicates the cumulative number of DEPs exceeding that threshold (FDR < 0.05). The dashed red line marks the applied threshold of |log_2_FC| = 0.5, yielding 1,966 DEPs. FC: fold change; DEPs: differentially expressed proteins.

**FIGURE S4 Principal component analysis (PCA) of salivary proteome data.**

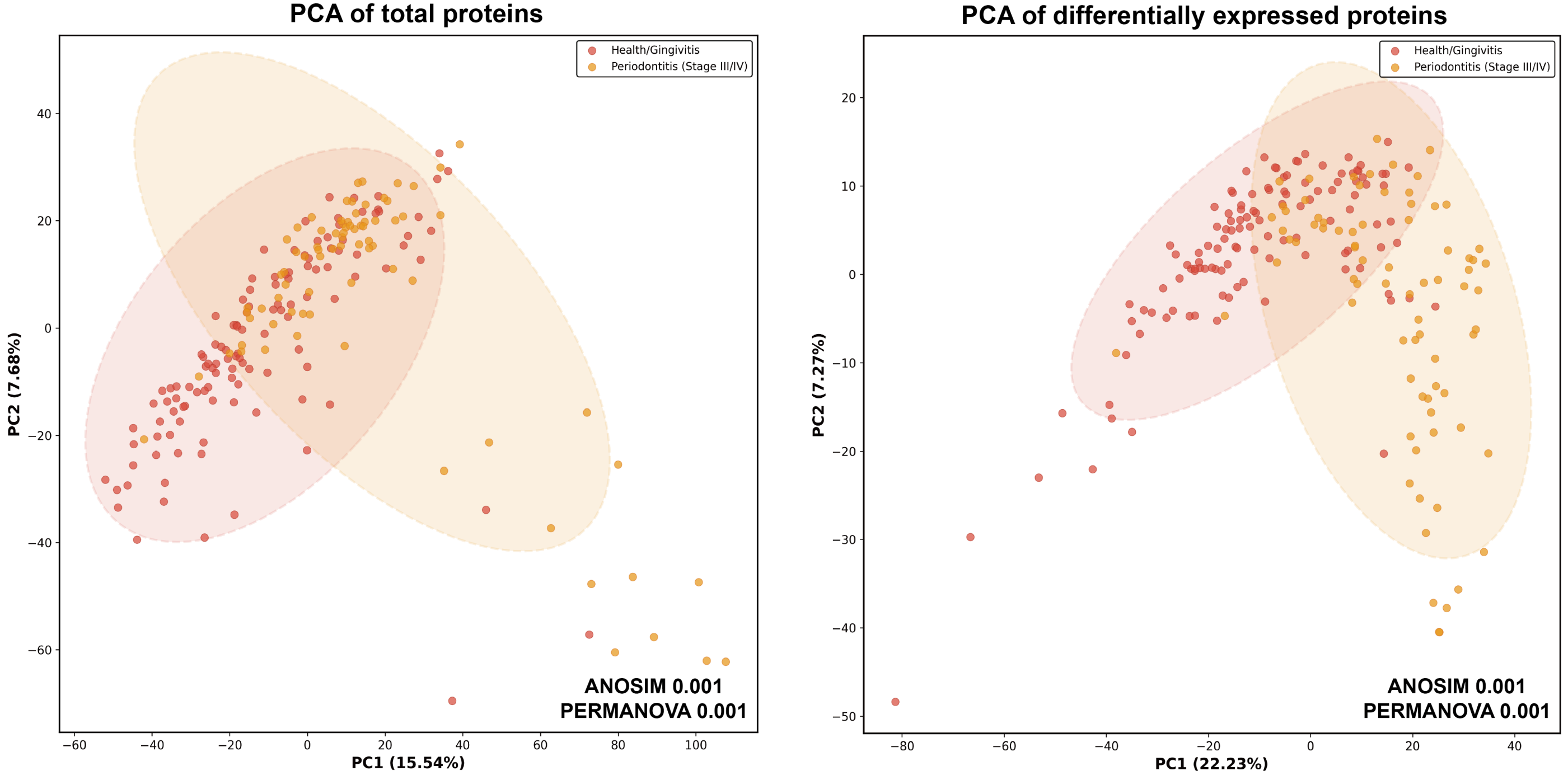

(Left) PCA of all quantified proteins. (Right) PCA of DEPs only. Each dot represents one saliva sample; red: H+G group; orange: P3+P4 group. Shaded ellipses indicate 95% confidence intervals. The percentage of variance explained by each principal component is indicated on the respective axis. ANOSIM and PERMANOVA p = 0.001, indicating statistically significant group separation. DEPs: differentially expressed proteins; PCA: principal component analysis; ANOSIM: analysis of similarities; PERMANOVA: permutational multivariate analysis of variance.

**FIGURE S5 Heatmap of the 1,966 differentially expressed proteins.**

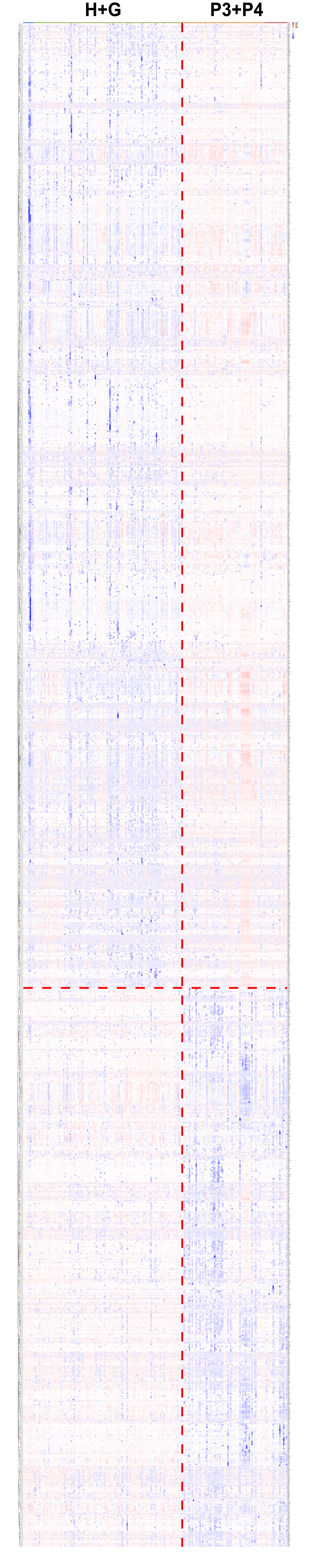

Heatmap displaying normalized protein expression profiles of the 1,966 differentially expressed proteins (rows) across all 199 saliva samples (columns), grouped by diagnosis (H+G, left; P3+P4, right). Rows and columns were independently clustered using unsupervised hierarchical clustering with Euclidean distance and complete linkage. Protein expression values were Z-score normalized by row prior to clustering (R package pheatmap). Red indicates higher relative expression; blue indicates lower relative expression. The vertical dashed red line demarcates the two sample groups; the horizontal dashed red line separates upregulated (above) from downregulated (below) proteins in the P3+P4 group. H+G: periodontal health and gingivitis; P3+P4: stage III/IV periodontitis; DEPs: differentially expressed proteins.

**FIGURE S6 Comparison of diagnostic performance across different biomarker panel sizes (2, 3, 5 proteins).**

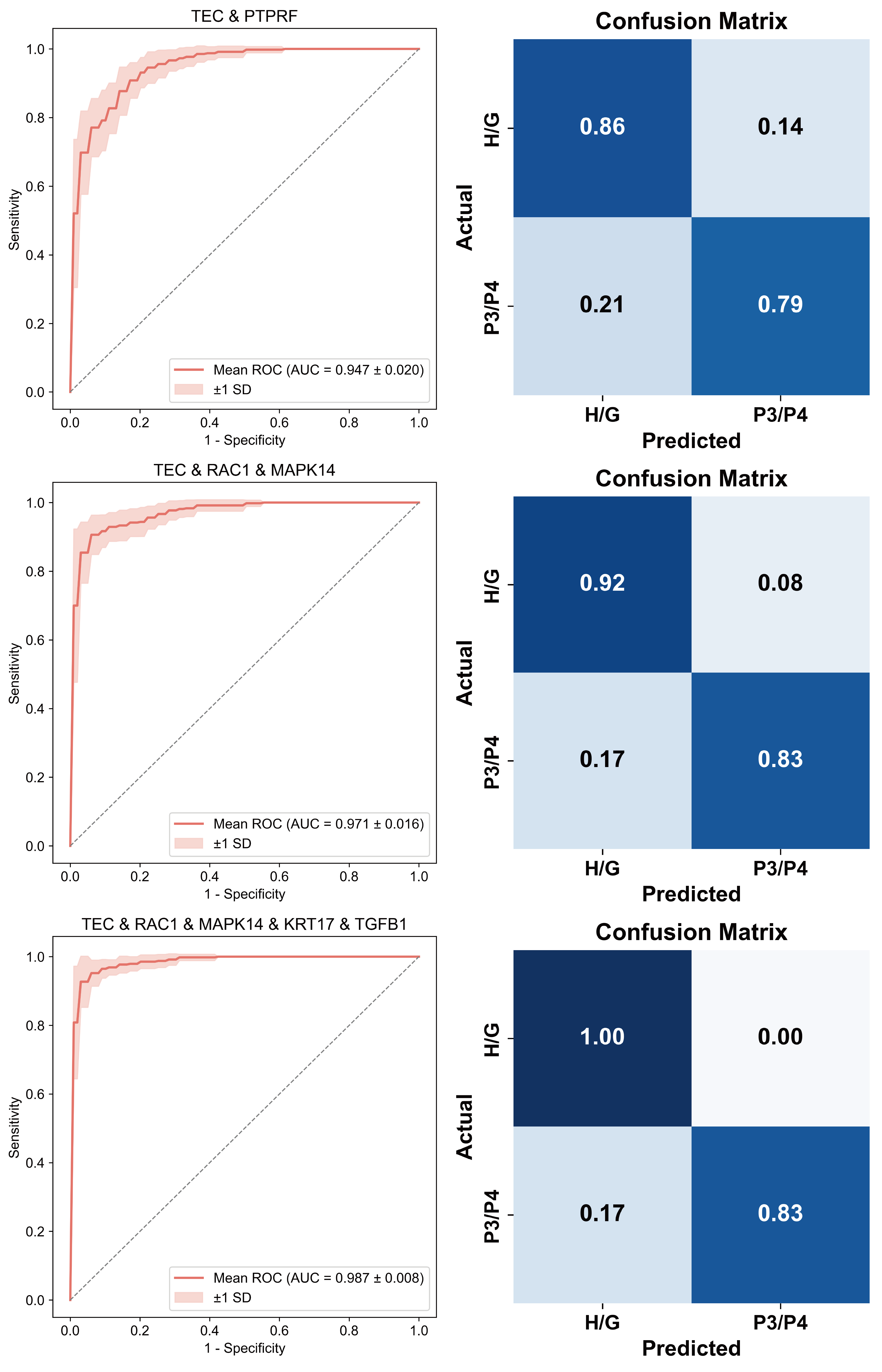

ROC curves (left) and confusion matrices (right) for biomarker panels of increasing size under 5-fold cross-validation. The 2-protein panel (TEC and PTPRF) achieved an AUC of 0.947 ± 0.020, with a specificity of 86% for the H+G group and a sensitivity of 79% for the P3+P4 group. The 3-protein panel (TEC, RAC1, and MAPK14) achieved an AUC of 0.971 ± 0.016, with a specificity of 92% and a sensitivity of 83%. The 5-protein panel (TEC, RAC1, MAPK14, KRT17, and TGFB1) achieved an AUC of 0.987 ± 0.008, with a specificity of 100% and a sensitivity of 83%. The shaded band represents ±1 SD. The ROC curve and confusion matrix for the selected 4-protein diagnostic panel are shown in Figure 5C–D. AUC: area under the curve; ROC: receiver operating characteristic; SD: standard deviation.

**FIGURE S7 Complete PPI network of the 1,966 differentially expressed proteins constructed using the STRING database.**

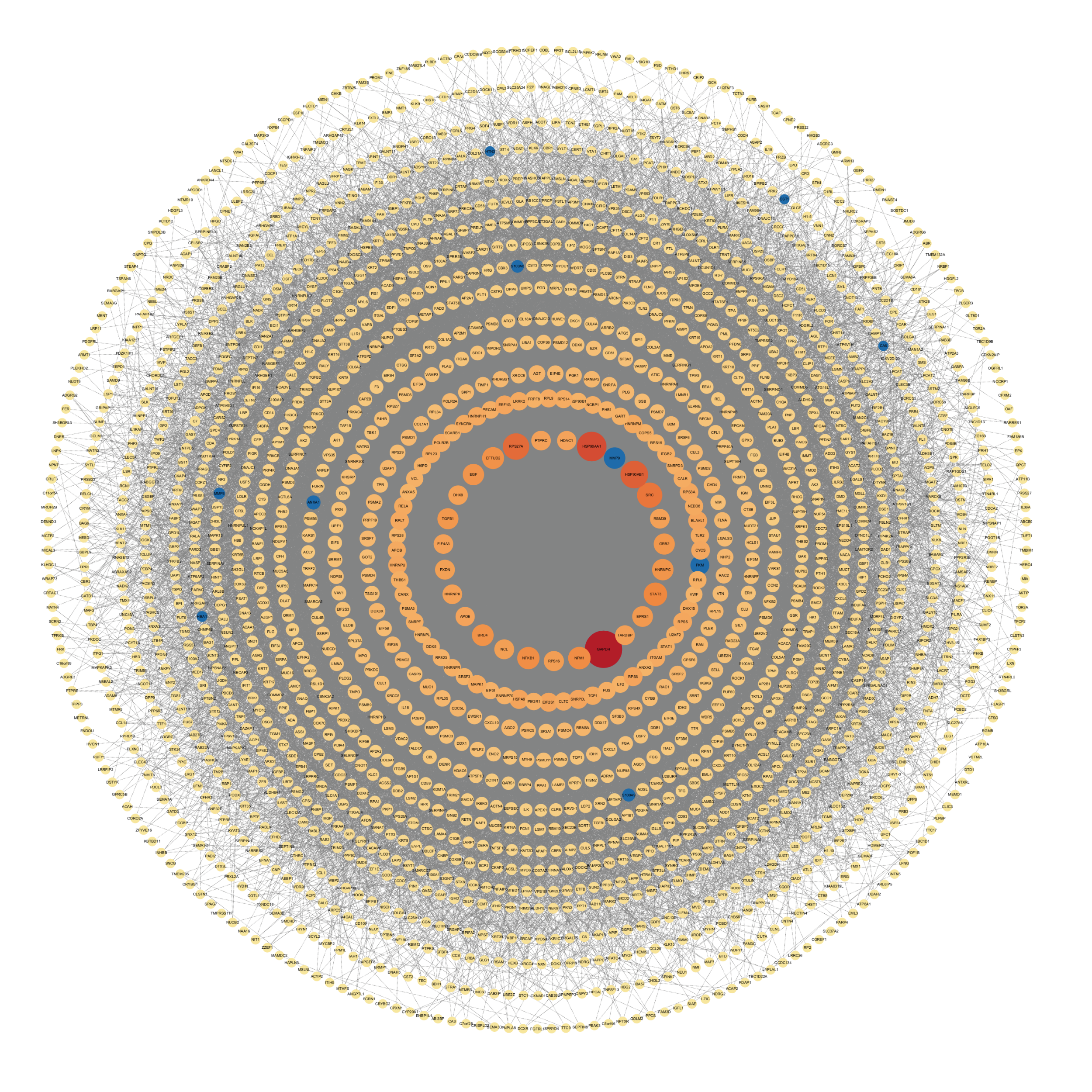
 PPI network of the 1,966 differentially expressed proteins constructed using the STRING database (confidence score ≥ 0.400), visualized using a degree-sorted concentric circle layout in Cytoscape. Each node represents a protein; proteins are arranged by degree — defined as the number of direct interaction partners within the network — with hub proteins of highest degree positioned in the innermost circle. Node color reflects connectivity degree, ranging from yellow (low) to red (high). Blue circles indicate proteins previously reported as periodontitis biomarkers in the literature. PPI: protein-protein interaction; DEPs: differentially expressed proteins.

**FIGURE S8 PPI network analysis exploring functional relationships between the four-protein biomarker panel (TEC, RAC1, MAPK14, KRT17) and the 274 DEPs identified from the external validation dataset.**

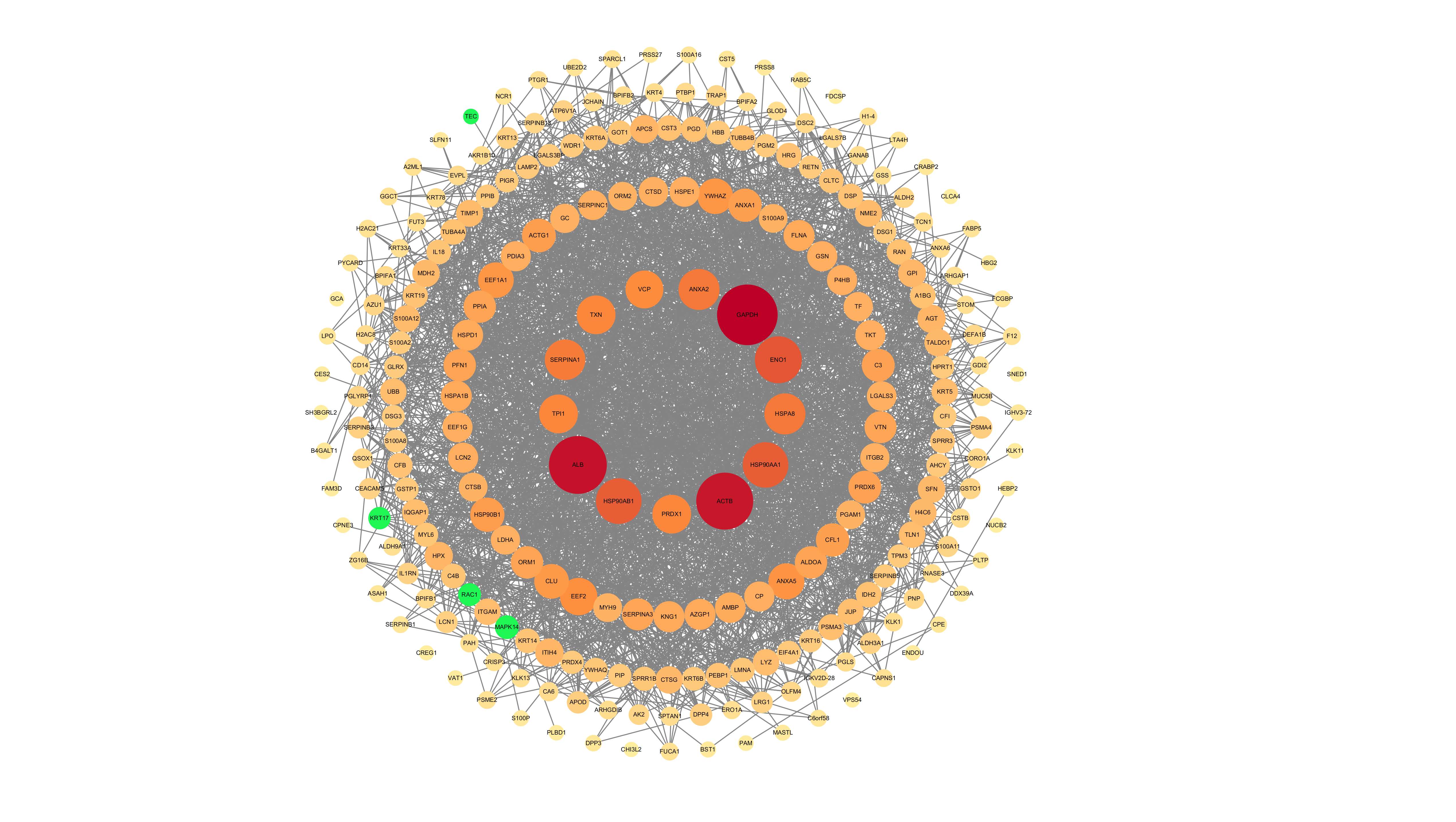

PPI network of the 274 differentially expressed proteins identified from the external validation dataset, constructed using the STRING database (confidence score ≥ 0.400), and visualized using a degree-sorted concentric circle layout in Cytoscape. Each node represents a protein; proteins are arranged by degree — defined as the number of direct interaction partners within the network — with hub proteins of highest degree positioned in the innermost circle. Node color reflects connectivity degree, ranging from yellow (low) to red (high). The four proteins of the diagnostic panel (TEC, RAC1, MAPK14, KRT17) are highlighted in green. PPI: protein-protein interaction; DEPs: differentially expressed proteins.
